# Risks and long-term prognosis of new-onset heart failure after *de novo* permanent pacemaker implantation: nationwide cohort study

**DOI:** 10.1101/2024.09.03.24313045

**Authors:** Young Jun Park, Sungjoo Lee, Sungjun Hong, Kyunga Kim, Juwon Kim, Ju Youn Kim, Kyoung-Min Park, Young Keun On, Seung-Jung Park

## Abstract

**Background:** Previous studies on pacemaker-associated heart failure (PaHF) have predominantly analyzed relatively small, single-center datasets, mainly focusing on incidence and predictors. However, the clinical implications of PaHF on mortality, particularly in relation to standard HF medications or upgrading to cardiac resynchronization therapy (CRT), has been underexplored.

**Methods:** Utilizing nationwide real-world data from the Korean National Health Insurance Service database, we analyzed 32,216 patients undergoing permanent pacemaker (PPM) implantation without preexisting HF between 2008 and 2019. The incidence, predictors, and mortality risk of PaFH were evaluated. To address potential immortal-time bias due to the time-dependent occurrence of PaHF, the time from the PPM implantation to the first diagnosis of PaHF was analyzed as a time-dependent covariate. For patients with PaHF, a propensity score-matched analysis was conducted based on CRT-upgrade status to explore the effect of CRT-upgrade on the risk of all-cause mortality.

**Results:** During the median 3.8-year follow-up period, PaHF and all-cause death occurred in 4170 (12.9%) and 6184 (19.2%) of the 32,216 PPM patients (42.3% male, mean age 70.6 years), respectively. PaHF development was closely associated with all-cause mortality, with a significantly higher mortality risk in the PaHF than in the non-PaHF group (hazard ratio [HR]=3.11, 95% confidence interval [CI]=2.93–3.32, P<0.001) after adjusting for immortal-time bias. The PaHF incidence and PaHF-associated mortality risk, although highest for the first six months post-PPM, did not disappear and increased again with follow-up time. In both the entire cohort (n=4170) and the propensity score-matched cohort (n=1685) of PaHF patients, CRT upgrade (HR=0.34, 95% CI=0.24–0.47, P<0.001), the use of beta-blockers (HR=0.75, 95% CI=0.61–0.93, P=0.010), and angiotensin receptor neprilysin inhibitor (ARNI) use (HR=0.28, 95% CI=0.14–0.54, P<0.001) were identified as potent protective factors against post-PaHF all-cause mortality.

**Conclusions:** PaHF development independently predicted post-PPM mortality, while upgrading to CRT and the use of beta-blockers or ARNI were identified as favorable prognostic factors for post-PaHF overall survival. Therefore, for PaHF patients, an immediate change into CRT or conduction system pacing, may be required along with optimal HF medications owing to the ongoing mortality risk.

## Introduction

Permanent pacemaker (PPM) implantation is the definitive treatment for patients with symptomatic bradyarrhythmia, such as sinus node dysfunction (SND) or atrioventricular block (AVB). However, pacing-induced left ventricular (LV) electromechanical dyssynchrony can lead to progressive LV dilatation, LV ejection fraction (EF) deterioration, and, eventually, clinical heart failure (HF), which can be specifically defined as pacemaker-associated HF (PaHF). ^1–4^

PaHF occurs in approximately 6–25% of patients with PPM 2–4 years after implantation. ^1,5–7^ Several PaHF risk factors have been reported, including older age, male, pre-existing LV systolic dysfunction, baseline left bundle branch block, right ventricular apical pacing, prolonged paced QRS duration, and a higher right ventricular (RV)-pacing percentage. ^1–3,5–8^ In addition, patients with PaHF had a significantly higher mortality rate than those without, ^1,6^ suggesting that a better understanding of PaHF is essential for improving the prognosis of this patient population. However, most previous studies on PaHF were single-center studies with relatively small sample sizes, primarily focusing on the incidence and predictors of PaHF. Moreover, the clinical impact of PaHF on mortality has only been evaluated in a limited number of studies. ^1,5,6^ In addition, the role of standard HF medications or upgrading to cardiac resynchronization therapy (CRT) devices on the prognosis of PaHF has not been thoroughly investigated in a large-scale registry.

Accordingly, we sought to investigate the incidence, predictors, and prognosis of PaHF using a nationwide database. Furthermore, we evaluated the effects of standard HF medications or CRT-upgrade on overall mortality in PaHF patients.

## Methods

### Data sources

This nationwide retrospective cohort study used data from the National Health Information Database (NHID) administered by the Korean National Health Insurance Service. The National Health Insurance Service is a mandatory, single payer social health insurance system that is managed by the Korean government and covers almost the entire (≥52 million) South Korean population. ^9^ The NHID contains comprehensive information on healthcare services, including demographics, diagnoses (based on the International Classification of Disease 10th Revision [ICD-10]), hospital visits and admissions, prescription drugs (based on Anatomical Therapeutic Chemical codes), procedures, medical device usage, and mortality data. It was established nationwide in 2000 and has been accessible to the public on the National Health Insurance Data Sharing Service homepage (http://nhiss.nhis.or.kr), primarily for academic or public policy purposes since 2009. ^10,11^ Favorable reliability of the diagnostic codes in the NHID has previously been reported for major cardiovascular or intractable diseases, such as hypertrophic cardiomyopathy (93% accuracy) or myocardial infarction (92% accuracy). ^9,12,13^ This study was approved by the Institutional Review Board (IRB) of Samsung Medical Center (IRB File No. 2019-05-075). The requirement of informed consent was waived as data are public and deidentified under confidentiality guidelines.

### Study design and population

Two retrospective cohorts were designed for a twofold purpose. The primary cohort consists of patients with *de novo* PPM implantation without preexisting HF to evaluate the incidence of PaHF and the all-cause mortality risk (PPM Cohort, Supplementary Figure 1). We identified 38,921 adult patients (≥18 years) who underwent *de novo* PPM implantation between January 1, 2008, and December 31, 2019, using the procedure and device codes for claims reimbursement (Supplementary Table 1). We excluded patients who had a previous history of

HF before PPM implantation (n=6,389). The previous history of HF was defined as hospitalization with HF code (I50.9) or prescription history of angiotensin receptor neprilysin inhibitor (ARNI) prior to PPM implantation. Additional exclusions were made for those who underwent reimplantation of a pulse generator and pacing leads following PPM system removal (n=892), or who died on the day of PPM implantation (n=6). Finally, the primary cohort contained 32,216 patients for analyses, including 4,170 patients who developed PaHF and 28,046 patients who did not, respectively.

The secondary cohort consisted only of 4,166 patients who developed PaHF in the primary cohort, excluding 4 who died on the day of PaHF diagnosis (PaHF Cohort, Supplementary Figure 1). The secondary cohort was further stratified into patients who were managed with CRT-upgrade (n=330) and those without (n=3,836) to explore the effect of CRT-upgrade on the risk of all-cause mortality. To emulate a randomized controlled trial, we further built a propensity-score (PS) matched cohort of PaHF patients with 1:4 ratio, including 316 and 1,139 patients with and without CRT-upgrade, respectively.

### Data acquisition

Demographic data such as age and sex were obtained. Comorbidities were identified based on ICD-10 codes from claims data between one year before the index date and the date of PaHF (Supplementary Table 2). The Charlson comorbidity index (CCI) that represents the overall comorbidity load at baseline was calculated based on CCI-related variables with their ICD-10 codes (Supplementary Table 3). We identified medication history for renin-angiotensin system (RAS) inhibitors including angiotensin-converting-enzyme inhibitors (ACEIs), angiotensin II receptor blockers (ARBs), and ARNI. Information on beta-blockers, mineralocorticoid receptor antagonists (MRAs), loop diuretics, thiazide, antiplatelet agents, and anticoagulants were also retrieved. Pacemaker type and CRT-upgrade were defined using ICD-10 codes for cardiac implantable electronic device and procedure (Supplementary Table 1).

### Study outcomes and follow-up

Two outcomes were considered in the analyses of the primary PPM cohort: the incidences of PaHF and all-cause death after *de novo* PPM implantation. First, PaHF was strictly defined as hospitalization with a newly assigned HF code (I50.9) following PPM implantation and ≥2 claims for HF medications among ACEIs/ARBs, beta-blockers, or MRAs. In addition, PPM patients who received CRT-upgrade or ARNI treatment were considered to meet the PaHF definition, regardless of HF code assignment or utilization of other HF medications. A broad PaHF definition was further considered to include all patients with a newly assigned HF code. Details of PaHF definitions are summarized in Supplementary Table 4. All main analyses were conducted based on the strict PaHF definition, whereas sensitivity analyses were based on the broad definition. Time-to-PaHF was defined as the time from the date of PPM implantation to the first diagnosis of PaHF during the follow-up. It was censored at the time of death, at the time of new HF-related events developing prior to PaHF diagnosis due to causes other than PaHF, such as myocardial infarction, myocarditis, alcoholic cardiomyopathy, cardiac sarcoidosis, and cardiac amyloidosis (Supplementary Table 5), or at the time of last follow-up (December 31, 2019), whichever came first. Second, post-PPM follow-up duration for all-cause mortality was defined as the time from the date of PPM implantation to the all-cause death or the date of last follow-up, whichever came first.

The outcome of the secondary PaHF cohort was all-cause death. Post-PaHF follow-up duration for all-cause mortality was defined as the time from the first diagnosis of PaHF to the all-cause death or the date of last follow-up, whichever came first.

### Statistical analyses

Baseline characteristics were described as means with standard deviations for continuous variables, and as counts with percentages for categorical variables. Continuous variables were compared using a Student’s t-test or Mann–Whitney rank-sum test, whereas categorical variables were compared using the χ2 test or Fisher exact test, as appropriate. Incidence rates of PaHF and all-cause death were calculated as events per 100 patient-years (PYs) with exact Poisson 95 % confidence intervals (CIs).

The cumulative incidence rate of PaHF was estimated using the Kaplan-Meier (K-M) method. In addition, the instantaneous incidence rate of PaHF was calculated using a nonparametric smoothing method based on B-splines and a generalized linear mixed model across the entire follow-up period. ^14^ Multivariable Cox proportional hazards (PHs) regression analysis was performed and HRs with 95% CIs were estimated to explore risk factors associated with the occurrence of PaHF.

In the analysis of all-cause mortality after *de novo* PPM implantation, PaHF was considered as a primary exposure. Because PaHF exposure was determined during the follow-up period rather than at baseline, there exists a potential immortal-time bias when comparing the mortality risk according to PaHF. Immortal time refers to the duration wherein the outcome could not have occurred until the exposure was determined in observational studies^15^, and was the time from the PPM implantation to the first diagnosis of PaHF in this study (Supplementary Figure 2). To account for the potential immortal-time bias, PaHF was considered as a time-dependent covariate in the extended K-M estimation and multivariable Cox PH regression analysis. The extended K-M estimator updates the cohorts at each time of PaHF development by allowing the PaHF group to contribute risk to the non-PaHF group before they experience PaHF. ^16^ In addition, multivariable Cox regression analysis with cubic spline functions was performed to investigate the time-varying effect of time-dependent PaHF occurrence on all-cause mortality across the entire follow-up period. ^17^ To evaluate a possible modification on the association between the PaHF and mortality, we conducted subgroup analyses stratified by age (<65, 65–75, and >75 years), sex, comorbidities, pacemaker type (single-vs. dual-chamber), pacing indication (AVB vs. SND), and medication (user vs. non-user). The age stratification was determined based on studies and recent guidelines that have developed or recommended risk stratification schemes for predicting stroke and thromboembolism in patients with atrial fibrillation (AF). ^18–20^ The potential modification was tested by assessing the interaction term of each subgrouping variable with PaHF.

In the secondary PaHF cohort and also in its PS-matched cohort, post-PaHF all-cause mortality was compared between patient groups treated with and without CRT-upgrade (Supplementary Figure 1). PS was defined as the probability of receiving CRT-upgrade, and estimated using a multivariable binary logistic regression model with potential confounders. PaHF patients with CRT-upgrade were matched to those without at a 1:4 ratio by greedy matching without replacement using a caliper width equal to 0.25 times the pooled standard deviation of the logit of the PS. Standardized mean differences (SMDs) were used for balance diagnostics, and absolute values of SMD > 0.1 were considered as significant sign of imbalance. Overall survival curves were estimated by the K-M method, and were compared between the CRT-upgrade and non-upgrade groups using log-rank and stratified log-rank tests in the entire and the PS-matched PaHF cohorts, respectively. Multivariable Cox PH models were used to identify the associations between the CRT-upgrade status and all-cause mortality in both cohorts, and the robust standard errors were estimated by considering matched pairs as clusters in the matched cohort. To account for a potential violation against PH assumption, a sensitivity analysis was conducted using the extended K-M method and multivariable Cox regression with CRT-upgrade considered as a time-dependent covariate.

For our multivariable analysis, we carefully selected variables that have been previously investigated as potential risk factors or confounders for HF or mortality in patients with PPM or cardiovascular diseases. These variables include age, ^5,6, 21, 22^ sex, ^4–7,21–23^ diabetes mellitus (DM), ^4,5,24^ hypertension, ^4,6^ coronary artery disease, ^4,6,21^ peripheral artery disease, ^25^ chronic kidney disease including end stage renal disease (CKD/ESRD), ^5,21,26^ valvular heart disease, ^27^ AF, ^8,19,20,27,28^ chronic obstructive pulmonary disease, ^6,27^ pacing indication, ^5,22,29–31^ CRT-upgrade, ^2,32^ RAS inhibitors, ^7,32,33^ beta blockers, ^7,32,33^ MRAs. ^7,32,33^ To mitigate the risk of overfitting, we included covariates with P values < 0.05 from univariable analyses and clinically relevant variables (such as age and sex) in our multivariable analyses. Additionally, we performed supplementary multivariable analyses, incorporating variables in a non-parsimonious manner within both the primary and secondary cohorts to minimize the risk of inadvertently excluding potentially important variables from the multivariable analysis.

Multicollinearity was assessed by variance inflation factor whose value greater than 4 was considered to indicate non-negligible collinearity. In multivariable time-to-event analyses, the adjusted HRs were reported with 95% CIs. The PH assumption was tested based on the scaled Schoenfeld residuals. Statistical significance was considered with two-sided P values <0.05. Statistical analyses were performed using SAS version 9.3 (SAS Institute, Cary, NC, USA) and R version 4.1.0 (The R Foundation, www.R-project.org).

## Results

### Baseline characteristics of PPM cohort

Among 32,216 PPM patients in the primary cohort, 13,632 (42.3%), 20,246 (63.4%), and 27,073 (84.0%) had male sex, AVB, and dual-chamber PPMs, respectively. The mean age was 70.6±12.1 years. Details of baseline characteristics of PPM cohort are summarized in Table 1. The median follow-up duration was 3.8 (interquartile range, 1.7−6.7) years.

**Table 1.** Baseline characteristics of the PPM cohort.

| Variables | Overall<br>(n = 32 216) | PaHF<br>(n = 4170) | No PaHF<br>(n = 28 046) | aSMD | *P* value |
| --- | --- | --- | --- | --- | --- |
| *Demographics and medical history* |  |  |  |  |  |
| Age, years | 70.6 ± 12.1 | 71.9 ± 11.2 | 70.4 ± 12.2 | 0.128 | <.001 |
| Age ≥ 65 years | 24 033 (74.6) | 3314 (79.5) | 20 719 (73.9) | 0.133 | <.001 |
| Male | 13 632 (42.3) | 1815 (43.5) | 11 817 (42.1) | 0.028 | 0.093 |
| Diabetes | 10 291 (31.9) | 1578 (37.8) | 8713 (31.1) | 0.143 | <.001 |
| Hypertension | 24 711 (76.7) | 3557 (85.3) | 21 154 (75.4) | 0.250 | <.001 |
| Coronary artery disease | 13 003 (40.4) | 2170 (52.0) | 10 833 (38.6) | 0.272 | <.001 |
| Peripheral artery disease | 7453 (23.1) | 1015 (24.3) | 6438 (23.0) | 0.033 | 0.050 |
| CKD/ESRD | 2263 (7.0) | 390 (9.4) | 1873 (6.7) | 0.099 | <.001 |
| Valvular heart disease | 2976 (9.2) | 631 (15.1) | 2345 (8.4) | 0.211 | <.001 |
| Atrial fibrillation | 7297 (22.7) | 1169 (28.0) | 6128 (21.8) | 0.143 | <.001 |
| COPD | 6294 (19.5) | 1007 (24.1) | 5287 (18.9) | 0.129 | <.001 |
| CCI ≥ 3 unit | 19 612 (60.9) | 2885 (69.2) | 16 727 (59.6) | 0.200 | <.001 |
| *Pacemaker-related variables* |  |  |  |  |  |
| AV block | 20 246 (63.4) | 2502 (60.0) | 17 744 (63.3) | 0.068 | <.001 |
| Sinus node dysfunction | 11 675 (36.6) | 1631 (39.1) | 10 044 (35.8) |  |  |
| Dual chamber | 27 073 (84.0) | 3136 (75.2) | 23 937 (85.3) | 0.262 | <.001 |
| *Medications* |  |  |  |  |  |
| ACEIs or ARBs | 21 490 (66.7) | 3309 (79.4) | 18 181 (64.8) | 0.328 | <.001 |
| Beta blockers | 13 905 (43.2) | 2390 (57.3) | 11 515 (41.1) | 0.330 | <.001 |
| MRAs | 5274 (16.4) | 1222 (29.3) | 4072 (14.5) | 0.363 | <.001 |
| Loop diuretics | 12 611 (39.1) | 2389 (57.3) | 10 222 (36.4) | 0.427 | <.001 |
| Thiazide | 6200 (19.2) | 1038 (24.9) | 5162 (18.4) | 0.158 | <.001 |
| Antiplatelet agents | 19 301 (59.9) | 2990 (71.7) | 16311 (58.2) | 0.287 | <.001 |
| Anticoagulants | 6382 (19.8) | 1127 (27.0) | 5255 (18.7) | 0.198 | <.001 |

Values are expressed as means ± standard deviations or n (%).
Abbreviations: PPM, permanent pacemaker; PaHF, pacemaker-associated heart failure; aSMD, absolute standardised mean difference; CKD, chronic kidney disease; ESRD, end-stage renal disease; COPD, chronic obstructive pulmonary disease; CCI, Charlson comorbidity index; AV block, atrioventricular block; ACEIs, angiotensin-converting enzyme inhibitors; ARBs, angiotensin II receptor antagonists; MRAs, mineralocorticoid receptor antagonists.

### PaHF incidence and risk factors

New-onset PaHF developed in 4,170 (12.9%) of 32,216 PPM patients with an incidence rate of 3.3 per 100 PYs (95% CI 3.2−3.4) (Supplementary Figures 1 and 3). The mean time-to-PaHF was 3.0±2.8 years. When the broad PaHF definition was used, PaHF was observed in 6,118 (19.0%) patients with much higher incidence rate of 4.9 per 100 PYs (95% CI 4.8−5.0).

Patients in the PaHF group exhibited worse baseline features for most variables: more advanced age, a higher proportion of comorbidities, and more frequent use of medications (Table 1). The multivariable Cox regression analysis identified age (as continuous or dichotomous variable), male, and various comorbidities as independent risk factors of PaHF development (Table 2); diabetes mellitus (DM), hypertension, coronary heart disease, chronic kidney disease including end stage renal disease (CKD/ESRD), valvular heart disease, AF, and chronic obstructive pulmonary disease were significantly associated with PaHF occurrence. In contrast, AVB (vs. SND) and peripheral artery diseases did not significantly affect PaHF occurrence after PPM implantation.

**Table 2.**
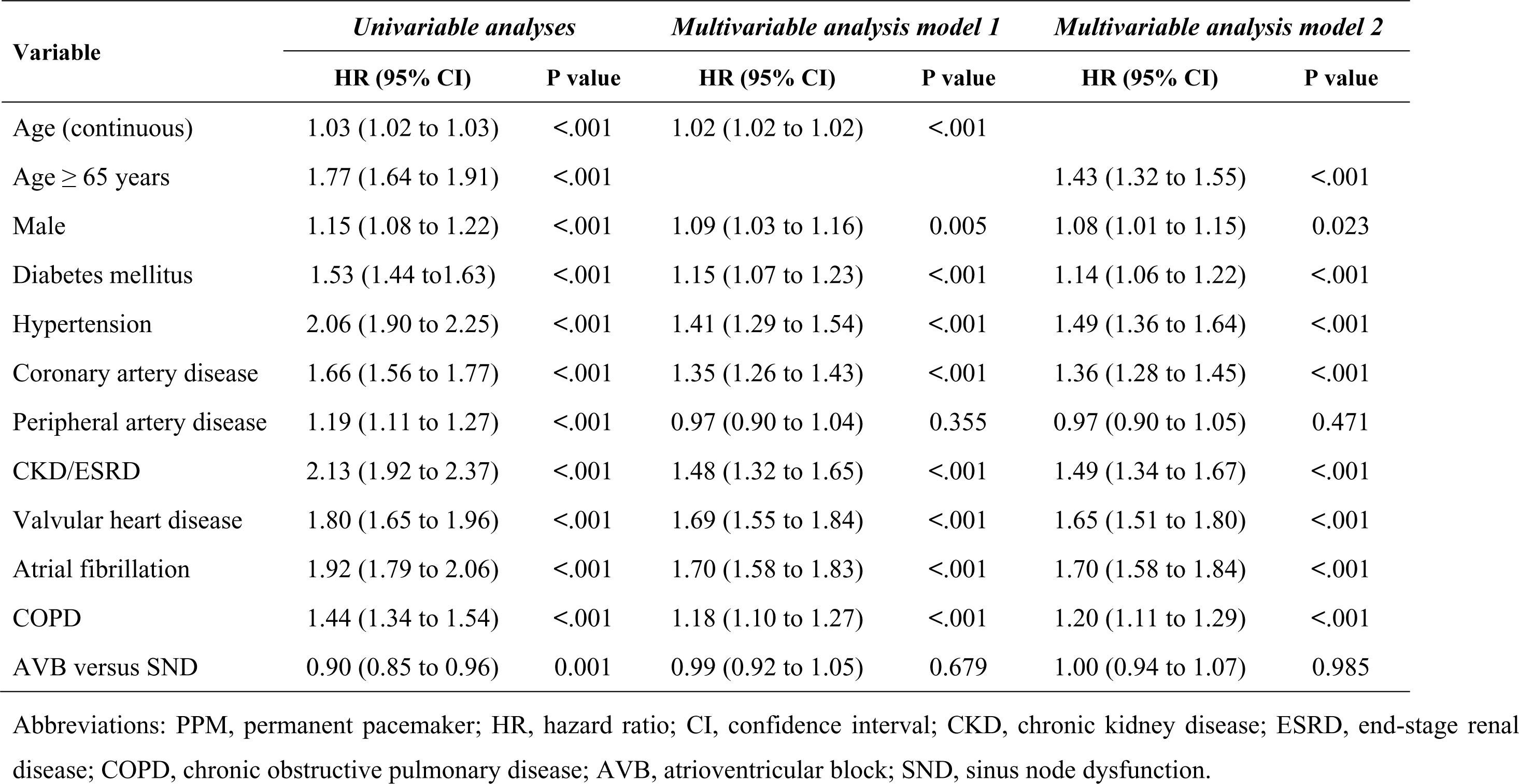
Independent risk factors of pacemaker-associated heart failure in the PPM cohort.

### Risk of all-cause mortality according to the development of PaHF

During the study period, all-cause deaths occurred in 6,184 (19.2%) PPM patients. Patients with PaHF had higher incidence rate of all-cause death than those without; 6.2 (95% CI 5.9−6.5) vs. 4.0 (3.9−4.1) per 100 PYs, P <0.001. After adjusting for immortal-time bias, the extended K-M curves also demonstrated worse prognosis of the PaHF compared to non-PaHF groups (Figure 1). On multivariable Cox PH regression analysis, PaHF development was identified as an independent risk factor of post-PPM all-cause mortality (HR 3.11, 95% CI 2.93−3.32, P <0.001), with adjustment for immortal-time bias as well as potential confounders, including age, sex, DM, hypertension, coronary artery disease, peripheral artery disease, CKD/ESRD, valvular heart disease, AF, chronic obstructive pulmonary disease, pacing indication, CCI, types of PPM, and medications (Supplementary Table 6). In most subgroup analyses, patients with PaHF consistently had higher mortality risk than those without (Figure 2).

**Figure 1.**
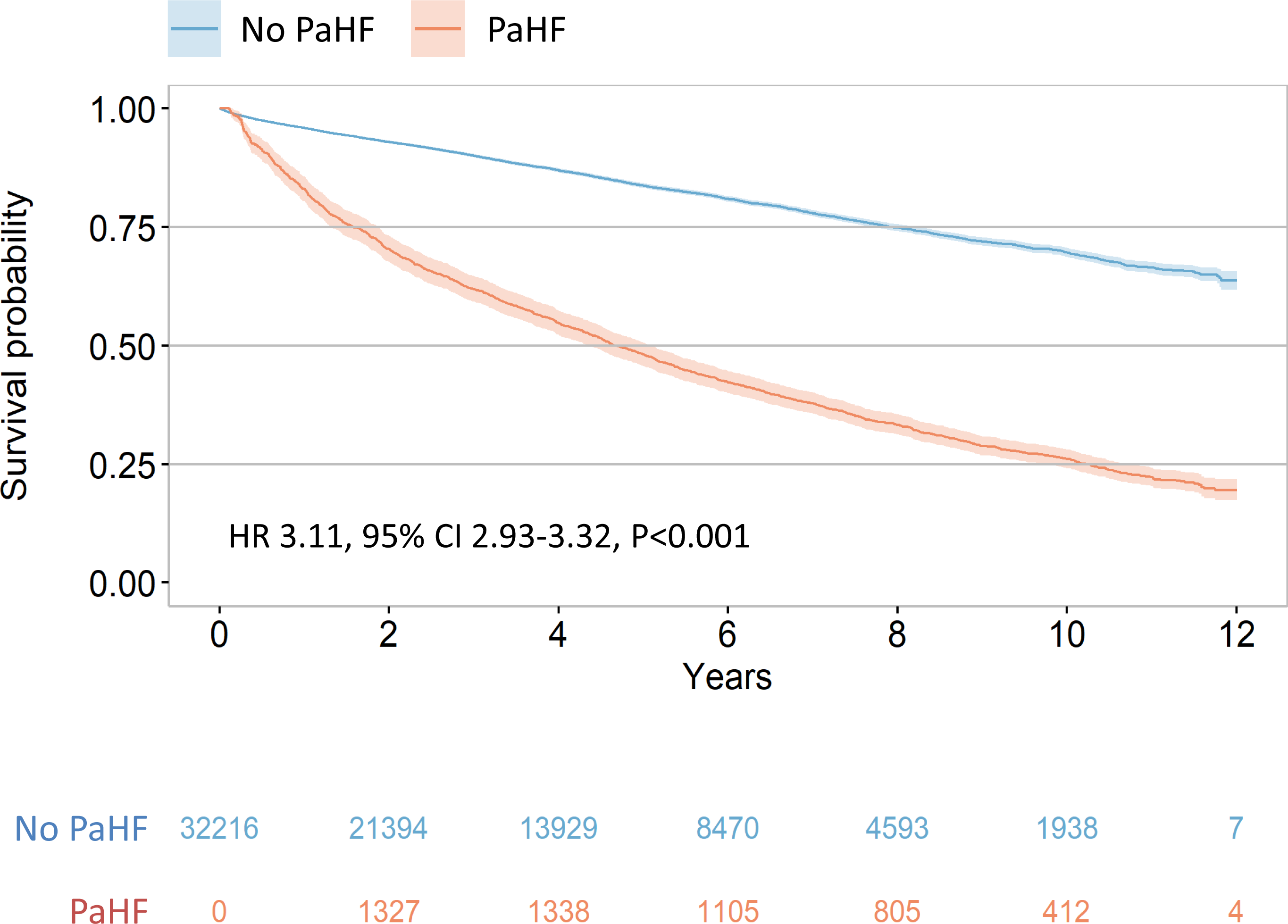
Extended Kaplan–Meier survival curves for all-cause death. Overall survival of patients with PaHF was significantly reduced compared to those without, even adjusted for time from PPM implant to PaHF diagnosis as a time-dependent covariate. Hazard ratio (HR) of patients with PaHF relative to those without, along with 95% confidence interval (CI) and P value for the difference between the two curves, were derived from the corresponding adjusted Cox regression analysis. Abbreviations: PaHF, pacemaker-associated heart failure; PPM, permanent pacemaker.

**Figure 2.**
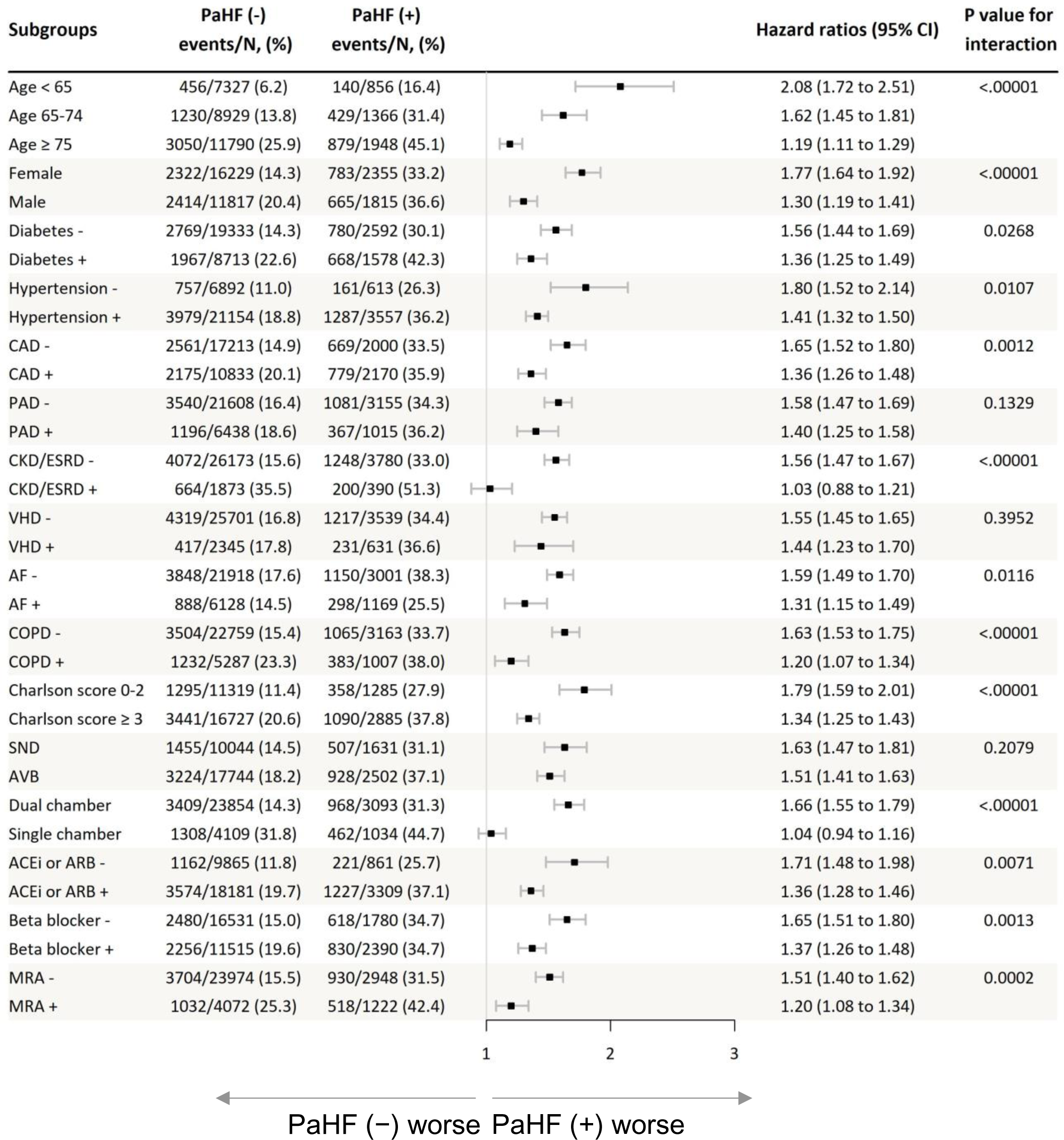
Forest plot of association between PaHF and all-cause mortality by subgroups. Patients with PaHF were consistently associated with higher mortality than those without, regardless of age, sex, comorbidities, or use of HF medications. Abbreviations: PaHF, pacemaker-associated heart failure; CI, confidence interval; CAD, coronary artery disease; PAD, peripheral artery disease; CKD, chronic kidney disease; ESRD, end-stage renal disease; VHD, valvular heart disease; AF, atrial fibrillation; COPD, chronic obstructive pulmonary disease; SND, sinus node dysfunction; AVB, atrioventricular block; ACEI, angiotensin-converting enzyme inhibitor; ARB, angiotensin receptor blocker; MRA, mineralocorticoid receptor antagonist.

### Time courses of PaHF incidence and PaHF-associated mortality

The instantaneous incidence rates of strictly- and broadly-defined PaHFs were highest during the first 6 months following PPM implantation (Figure 3A). However, they remained above zero during the entire follow-up period, making L-shaped curves. Analysis of the time-varying effect of PaHF on all-cause mortality, revealed that the PaHF-associated mortality risk was also highest in the first 6 months, then gradually decreased reaching its nadir around 5 years post-PPM (Figure 3B). Thereafter it began to rise up again with follow-up time, creating a U-shaped curve. Overall, the risk of PaHF-associated mortality remained significantly throughout the entire follow-up period.

**Figure 3.**
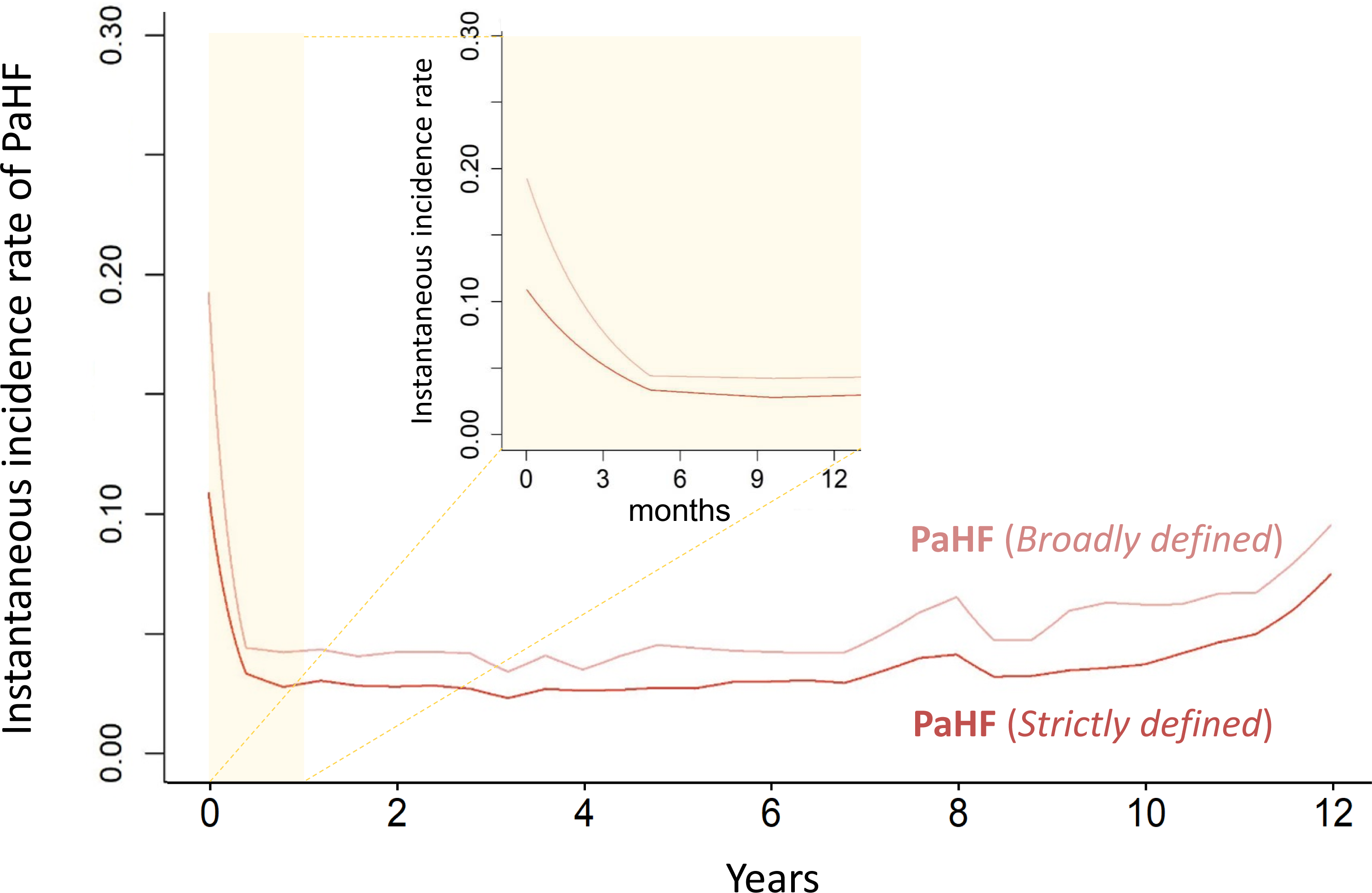
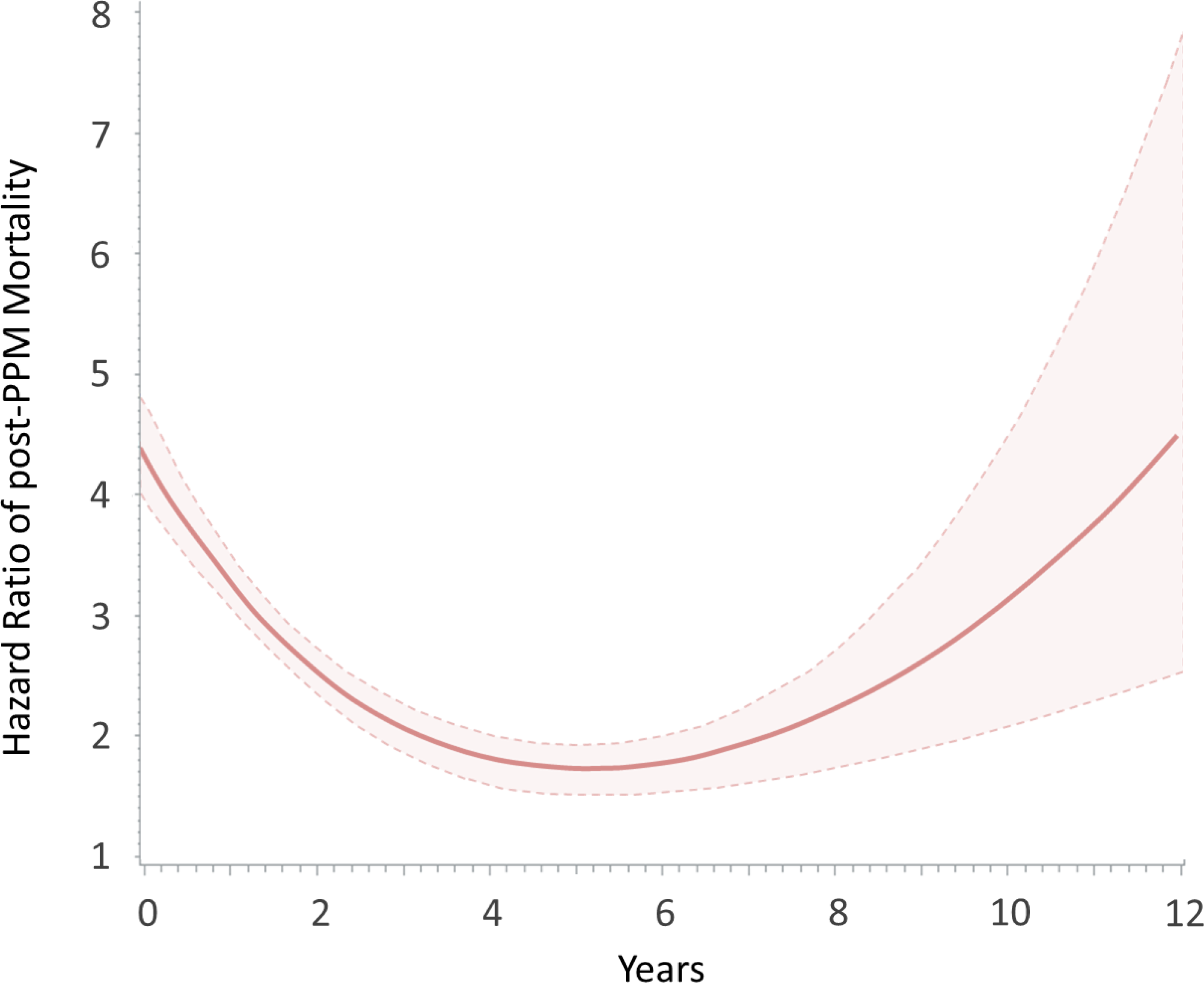
Time-varying instantaneous incidence rates of PaHF occurrence (A) and hazard ratio of PaHF on all-cause mortality (B) throughout the entire follow-up period. Enlarged graphs depicting the first year after PPM implant were overlayed for clarity (A). The hazard ratios of post-PPM mortality by cubic spline Cox regression analysis were invariably greater in the PaHF group than in the non-PaHF group throughout the entire follow-up period. The solid and dashed lines represent the hazard ratio and its 95% confidence interval, respectively (B). Abbreviations: PaHF, pacemaker-associated heart failure; PPM, permanent pacemaker.

### Prognosis of patients with PaHF according to CRT-upgrade

Table 3 displayed baseline characteristics at the time of PaHF diagnosis for the entire and PS-matched cohorts of PaHF patients. In the PS-matched cohort, most baseline characteristics were well balanced between the two treatment groups, except for age ≥65 years (absolute SMD, 0.131) and use of ARNI (0.212) or loop diuretics (0.219).

**Table 3.** Baseline characteristics of PaHF cohorts.

| Variables | Entire PaHF cohort |  |  | PS-matched PaHF cohort |  |  |
| --- | --- | --- | --- | --- | --- | --- |
|  | CRT-upgrade *<br>(n = 330) | No CRT-upgrade *<br>(n = 3836) | aSMD | CRT-upgrade *<br>(n = 316) | No CRT-upgrade *<br>(n = 1139) | aSMD |
| *Demographics and medical history* |  |  |  |  |  |  |
| Age, years | 67.5 ± 13.7 | 75.6 ± 10.5 | 0.670 | 67.7 ± 13.4 | 70.3 ± 12.4 | 0.196 |
| Age ≥ 65 years | 215 (65.2) | 3313 (86.4) | 0.511 | 207 (65.5) | 817 (71.7) | 0.131 |
| Male | 155 (47.0) | 1658 (43.2) | 0.075 | 149 (47.2) | 524 (46.0) | 0.022 |
| Diabetes | 89 (27.0) | 1270 (33.1) | 0.134 | 84 (26.6) | 342 (30.0) | 0.078 |
| Hypertension | 247 (74.8) | 2944 (76.7) | 0.044 | 236 (74.7) | 895 (78.6) | 0.089 |
| Coronary artery disease | 183 (55.5) | 2725 (71.0) | 0.328 | 174 (55.1) | 651 (57.2) | 0.042 |
| Peripheral artery disease | 113 (34.2) | 1573 (41.0) | 0.140 | 111 (35.1) | 418 (36.7) | 0.033 |
| CKD/ESRD | 39 (11.8) | 720 (18.8) | 0.194 | 39 (12.3) | 168 (14.7) | 0.073 |
| Valvular heart disease | 60 (18.2) | 742 (19.3) | 0.030 | 54 (17.1) | 176 (15.5) | 0.043 |
| Atrial fibrillation | 116 (35.2) | 1881 (49.0) | 0.284 | 116 (36.7) | 439 (38.5) | 0.038 |
| COPD | 117 (35.5) | 1793 (46.7) | 0.231 | 111 (35.1) | 419 (36.8) | 0.035 |
| *Pacemaker-related variables* |  |  |  |  |  |  |
| AV block | 231 (70.0) | 2284 (59.5) | 0.255 | 226 (71.5) | 790 (69.4) | 0.048 |
| Sinus node disease | 90 (27.3) | 1524 (39.7) |  | 90 (28.5) | 349 (30.6) |  |
| Dual chamber | 289 (87.1) | 2843 (73.9) | 0.338 | 276 (87.3) | 987 (86.7) | 0.021 |
| *Medications* |  |  |  |  |  |  |
| RAS inhibitors<sup>†</sup> | 220 (66.7) | 2425 (63.2) | 0.072 | 209 (66.1) | 786 (69.0) | 0.061 |
| ARNI | 21 (6.4) | 296 (7.7) | 0.053 | 19 (6.0) | 126 (11.1) | 0.212 |
| Beta blockers | 151 (45.8) | 1964 (51.2) | 0.109 | 141 (44.6) | 523 (45.9) | 0.026 |
| MRAs | 106 (32.1) | 1121 (29.2) | 0.063 | 97 (30.7) | 359 (31.5) | 0.018 |
| Loop diuretics | 146 (44.2) | 2151 (56.1) | 0.238 | 135 (42.7) | 610 (53.6) | 0.219 |
| Thiazide | 17 (5.2) | 211 (5.5) | 0.016 | 17 (5.4) | 52 (4.6) | 0.036 |

Values were expressed as means ± standard deviations or n (%).
\*, CRT-upgrade and No CRT-upgrade represent patients who were treated with CRT-upgrade plus HF medications and those with HF medications alone without CRT-upgrade, respectively.
<sup>†</sup>, RAS inhibitors included ACEIs, ARBs, and ARNI.–
Abbreviations: PaHF, pacemaker-associated heart failure; PS, propensity score; CRT, cardiac resynchronisation therapy; aSMD, absolute standardised mean difference; CKD, chronic kidney disease; ESRD, end-stage renal disease; COPD, chronic obstructive pulmonary disease; AV block, atrioventricular block; ACEIs, angiotensin-converting enzyme inhibitors; ARBs, angiotensin II receptor antagonists; ARNI, angiotensin receptor neprilysin inhibitor; MRAs, mineralocorticoid receptor antagonists.

During the median follow-up period of 1.9 (interquartile range, 0.7−2.6) years, a lower post-PaHF all-cause mortality was observed for the CRT-upgrade group, compared to the non-upgrade group in the entire cohort (incidence rate 4.1 [95% CI 3.0−5.5] vs. 14.6 [13.8−15.3] per 100PYs, P <0.001) and in the PS-matched cohort (4.1 [3.0−5.5] vs. 11.4 [10.3−12.6] per 100PYs, P <0.001). The K-M curve analyses also indicated a better prognosis for PaHF patients with CRT-upgrade compared to those without (log-rank P value <0.001 for the entire cohort and stratified log-rank P value <0.001 for the matched cohort) (Figures 4A and 4B).

**Figure 4.**
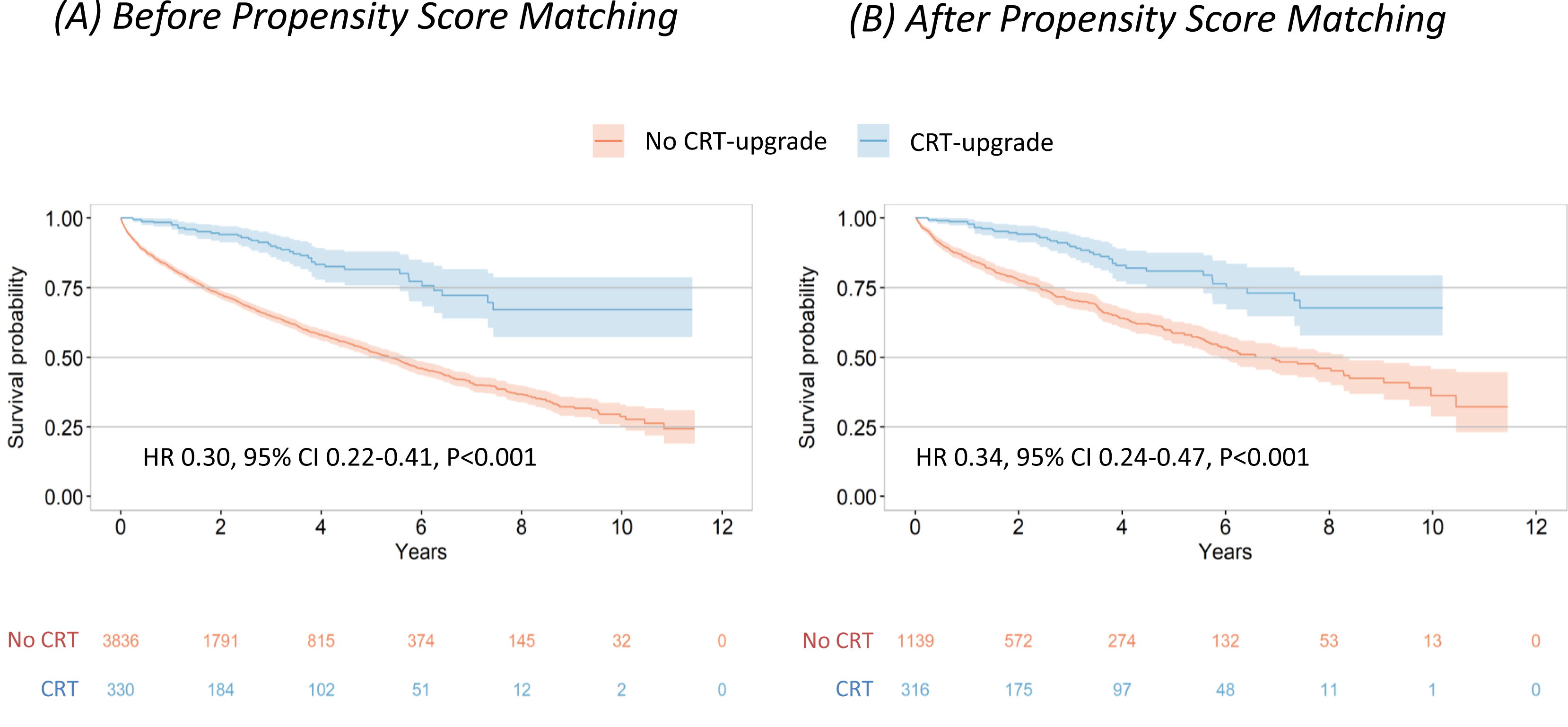
Kaplan–Meier survival curves for all-cause death in the secondary PaHF cohort. The long-term prognosis was significantly improved for patients who received CRT-upgrade in combination with medical treatment compared to those who received medical treatment alone without CRT-upgrade. Abbreviations: CRT, cardiac resynchronisation therapy; PaHF, pacemaker-associated heart failure.

Patient factors such as age (as a dichotomous or continuous variable), male, DM or CKD/ESRD were significantly associated with increased all-cause mortality, while treatment modalities including CRT-upgrade, RAS inhibitors, and beta-blockers were independent protective factors, with the lowest HR observed for CRT-upgrade in multivariable analysis model 1 (HR 0.34, 95% CI 0.24−0.47, P <0.001; Table 4 and Supplementary Table 7). However, when RAS inhibitors were broken down into ACEIs/ARBs and ARNI, only the use of ARNI was identified as a strong protective factor (HR 0.28, 95% CI 0.14−0.54, P <0.001; multivariable analysis model 2 in Table 4 and Supplementary Table 7).

**Table 4.**
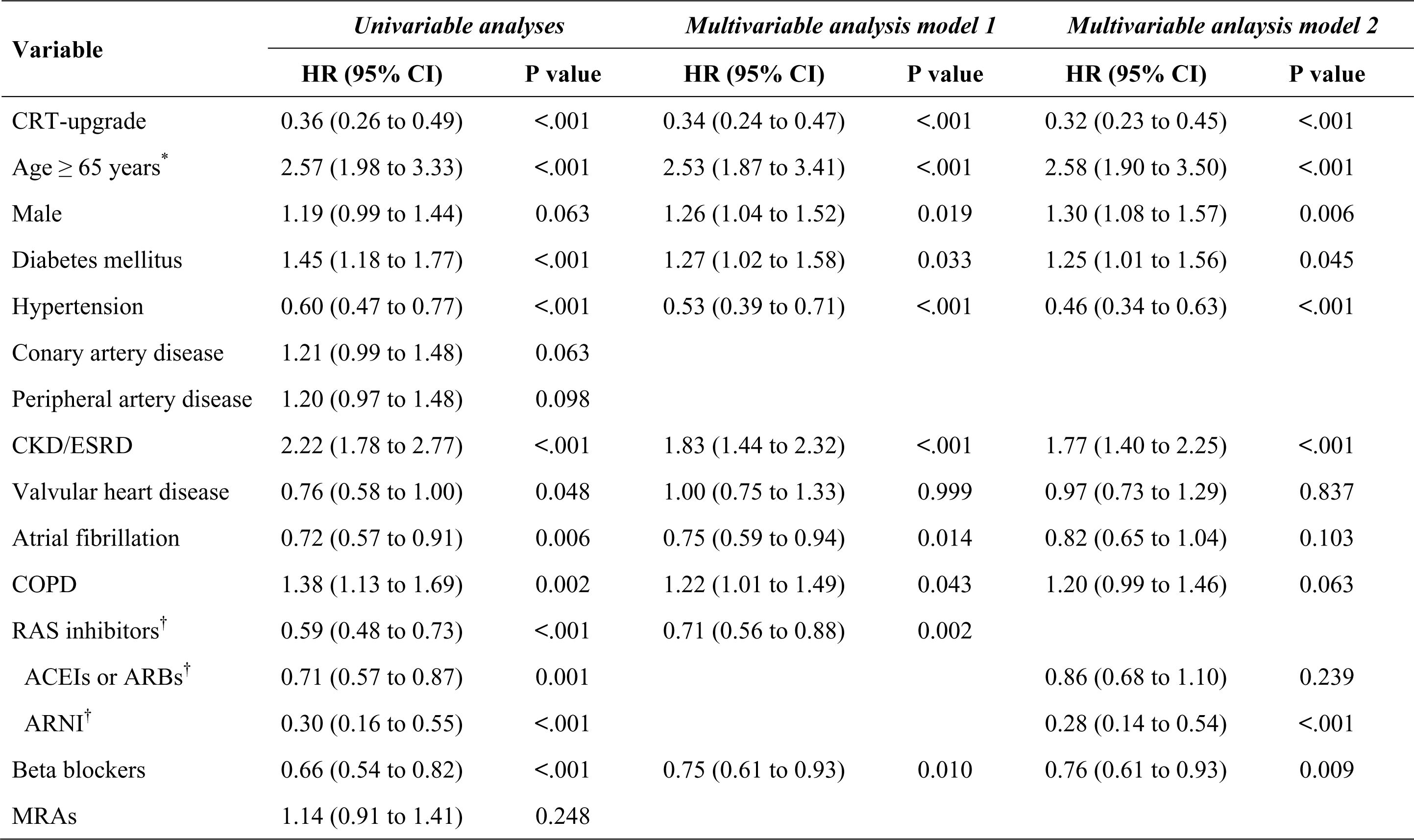
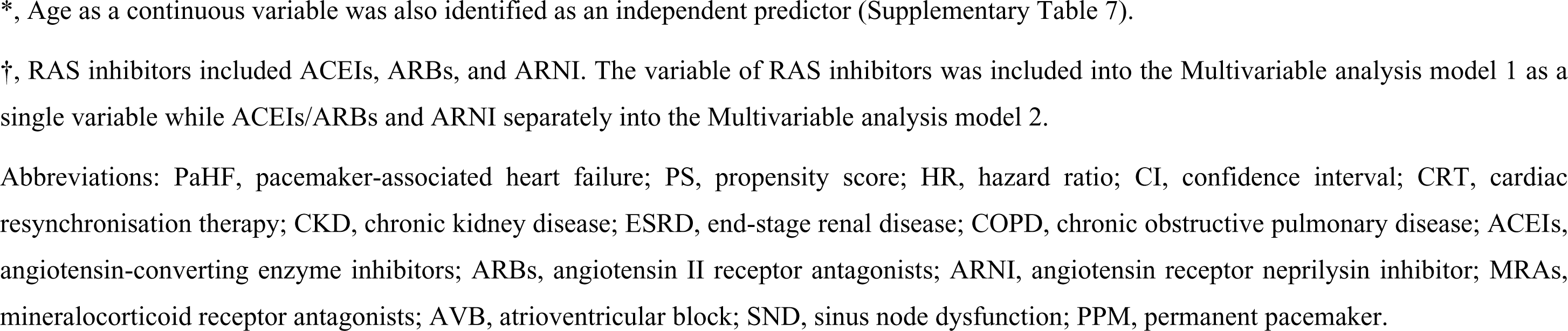
Independent risk factors of all-cause mortality in the propensity score-matched PaHF cohort.

| Variable | *Univariable analyses* |  | *Multivariable analysis model 1* |  | *Multivariable analysis model 2* |  |
| --- | --- | --- | --- | --- | --- | --- |
|  | HR (95% CI) | P value | HR (95% CI) | P value | HR (95% CI) | P value |
| CRT-upgrade | 0.36 (0.26 to 0.49) | <.001 | 0.34 (0.24 to 0.47) | <.001 | 0.32 (0.23 to 0.45) | <.001 |
| Age ≥ 65 years* | 2.57 (1.98 to 3.33) | <.001 | 2.53 (1.87 to 3.41) | <.001 | 2.58 (1.90 to 3.50) | <.001 |
| Male | 1.19 (0.99 to 1.44) | 0.063 | 1.26 (1.04 to 1.52) | 0.019 | 1.30 (1.08 to 1.57) | 0.006 |
| Diabetes mellitus | 1.45 (1.18 to 1.77) | <.001 | 1.27 (1.02 to 1.58) | 0.033 | 1.25 (1.01 to 1.56) | 0.045 |
| Hypertension | 0.60 (0.47 to 0.77) | <.001 | 0.53 (0.39 to 0.71) | <.001 | 0.46 (0.34 to 0.63) | <.001 |
| Coronary artery disease | 1.21 (0.99 to 1.48) | 0.063 |  |  |  |  |
| Peripheral artery disease | 1.20 (0.97 to 1.48) | 0.098 |  |  |  |  |
| CKD/ESRD | 2.22 (1.78 to 2.77) | <.001 | 1.83 (1.44 to 2.32) | <.001 | 1.77 (1.40 to 2.25) | <.001 |
| Valvular heart disease | 0.76 (0.58 to 1.00) | 0.048 | 1.00 (0.75 to 1.33) | 0.999 | 0.97 (0.73 to 1.29) | 0.837 |
| Atrial fibrillation | 0.72 (0.57 to 0.91) | 0.006 | 0.75 (0.59 to 0.94) | 0.014 | 0.82 (0.65 to 1.04) | 0.103 |
| COPD | 1.38 (1.13 to 1.69) | 0.002 | 1.22 (1.01 to 1.49) | 0.043 | 1.20 (0.99 to 1.46) | 0.063 |
| RAS inhibitors<sup>†</sup> | 0.59 (0.48 to 0.73) | <.001 | 0.71 (0.56 to 0.88) | 0.002 |  |  |
| ACEIs or ARBs<sup>†</sup> | 0.71 (0.57 to 0.87) | 0.001 |  |  | 0.86 (0.68 to 1.10) | 0.239 |
| ARNI<sup>†</sup> | 0.30 (0.16 to 0.55) | <.001 |  |  | 0.28 (0.14 to 0.54) | <.001 |
| Beta blockers | 0.66 (0.54 to 0.82) | <.001 | 0.75 (0.61 to 0.93) | 0.010 | 0.76 (0.61 to 0.93) | 0.009 |
| MRAs | 1.14 (0.91 to 1.41) | 0.248 |  |  |  |  |

\*, Age as a continuous variable was also identified as an independent predictor (Supplementary Table 7).
†, RAS inhibitors included ACEIs, ARBs, and ARNI. The variable of RAS inhibitors was included into the Multivariable analysis model 1 as a single variable while ACEIs/ARBs and ARNI separately into the Multivariable analysis model 2.
Abbreviations: PaHF, pacemaker-associated heart failure; PS, propensity score; HR, hazard ratio; CI, confidence interval; CRT, cardiac resynchronisation therapy; CKD, chronic kidney disease; ESRD, end-stage renal disease; COPD, chronic obstructive pulmonary disease; ACEIs, angiotensin-converting enzyme inhibitors; ARBs, angiotensin II receptor antagonists; ARNI, angiotensin receptor neprilysin inhibitor; MRAs, mineralocorticoid receptor antagonists; AVB, atrioventricular block; SND, sinus node dysfunction; PPM, permanent pacemaker.

### Sensitivity analyses

Results of sensitivity analyses based on the broad PaHF definition were consistent with the primary results based on the strict definition, demonstrating the PaHF group exhibited a significantly higher mortality than the non-PaHF group in the whole population (Supplementary Figures 4 and 5) and in various subgroup analyses (Supplementary Figure 6). Because the time from PaHF diagnosis to CRT-upgrade was very short (median 0.0, interquartile range 0.0−2.8 months), the immortal-time bias was not expected to be significant. Nonetheless, an additional sensitivity analysis was conducted to adjust for the immortal-time bias introduced by the PaHF-to-upgrade time. The results consistently indicated a significant protective effect of CRT-upgrade on the all-cause death (HR 0.45, 95% CI 0.32−0.63, P <0.001), compared to medical treatment alone without CRT-upgrade in patients with PaHF (Supplementary Table 8). In addition, we conducted additional multivariable analyses which incorporated variables in a non-parsimonious manner within both the primary and secondary cohorts. The overall results remained consistent (Supplementary Tables 6 and 9).

## Discussion

### Main findings and merits of this study

Our main findings were: (1) PaHF, when strictly or broadly defined, occurred in 12.9% and 19.0% of PPM patients, respectively, during long-term follow-up of 3.8 (interquartile range, 1.7−6.7) years; (2) the overall post-PPM mortality rate was 19.2%. PaHF development was an independent predictor for post-PPM all-cause death after adjusting for immortal-time bias and other potential confounders, and the risk was approximately three times higher in the PaHF group than in the non-PaHF group; (3) the PaHF incidence and PaHF-associated mortality rates were highest for the first six months post-PPM. However, the risk was likely persistent and increased again with follow-up time (Figures 3A and 3B); and (4) in the entire and propensity-matched cohorts of patients with PaHF, CRT-upgrade (HR 0.34), RAS inhibitors (HR 0.71), and beta blockers (HR 0.75) were identified as strong favorable prognostic factors for overall survival. However, when RAS inhibitors were categorized into ACEIs/ARBs and ARNI, a significant association with reduced mortality was observed only in ARNI, not in ACEIs/ARBs.

Our study has several merits compared to previous studies. First, this study was based on the two largest real-world cohorts to date: the PPM and PaHF cohorts, potentially enhancing the generalizability of our findings. Second, unlike previous studies^1,6^, we evaluated more rigorously the impact of PaHF on mortality risk, by addressing ‘the time-dependent occurrence of PaHF’ and ‘the time-dependent effect of PaHF on mortality’. Third, more importantly, our study is the first to investigate the effects of standard HF medical treatment, particularly the latest RAS inhibitor of ARNI, and upgrading to CRT devices on the prognosis of patients with PaHF, using a propensity score-matched nationwide cohort.

### PaHF incidence and predictors

During the median post-PPM follow-up period of 3.8 (interquartile range, 1.7−6.7) years, the PaHF incidence was 12.9% using the strict definition, which likely represented moderate to severe PaHF. However, the incidence increased to 19.0% using the broad definition, probably including mild forms of PaHF as well. Similar to our results, the PaHF incidence was 12.3% in a large single-center study where PaHF was strictly defined by including patients with moderate to severe LV systolic dysfunction. ^2^ However, a nationwide MarketScan database study reported that 25.8% of PPM patients developed PaHF, using a broader PaHF definition based only on the HF diagnosis code. ^28^

The independent predictors for PaHF identified in our study were also consistent with previous reports: advanced age (≥65 years), male, DM, CKD/ESRD, and AF. ^5–7,21,24,28^ However, contrary to our initial expectation, AVB was not associated with PaHF risk (Table 2). This discrepancy might be because pacing indications (e.g., AVB or SND) cannot reflect exact RV-pacing percentages. For example, in the DANPACE study, patients with SND who were presumed to have a low RV-pacing burden, had a much higher RV-pacing percentage of 85% with dual-chamber PPM implanted. ^29^ In contrast, in the IDEAL RVP study, the RV-pacing burden was below 40% in AVB patients who used PPM algorithms for minimizing RV-pacing. ^34^ This explanation may also be the reason why a large-scale German registry study showed no difference in the PaHF incidence (6% vs. 5.3%) or all-cause death (17% vs. 17%) rates between AVB and SND groups. ^5^

### Post-PPM mortality

Controversy remains regarding the mortality risk caused by chronic RV-pacing because PPM algorithms for minimizing the RV-pacing burden have failed to reduce all-cause mortality in previous randomized studies and a meta-analysis. ^5,22,29–31^ However, these unexpected results may be related to follow-up periods that were too short (e.g., mean duration of 2.5 years or less) to verify the mortality benefit of the algorithms. Indeed, in our data, the PPM-to-PaHF and PPM-to-death intervals were 3.0±2.8 years and 3.6±2.8 years on average, respectively. Moreover, the risk of post-PPM mortality rise up again approximately 5 years after PPM implant, making a U-shape curve over the entire follow-up period (Figure 3B and Supplementary Figure 5).

In line with previous reports, CKD, male, and advanced age (≥65 years) were significantly associated with increased mortality in patients with PaHF (Table 4). In CKD patients, several factors, including ventricular hypertrophy, myocardial fibrosis, and uremia, decelerate myocardial conduction velocity^35,36^, aggravating pacing-induced dyssynchrony. Furthermore, CKD and ESRD have been reported as independent predictors of PaHF and mortality in PPM patients. ^6,21,26^ Compared to women, men are more likely to have a larger heart size and greater myocardial mass, consequently requiring more time to activate the whole heart by single-site stimulation, and resulting in a more severe degrees of pacing-induced dyssynchrony and higher mortality. ^23,37^ Greater susceptibility to PaHF in male patients with larger hearts may be a counterpart phenomenon of better CRT responses in female patients with smaller hearts. ^38^ The specific role of DM on PaHF occurrence or PaHF-associated mortality has rarely been addressed. However, DM is frequently associated with advanced age, CKD/ESRD, and coronary artery disease, which could potentially impact mortality risk. ^24,39^

### PaHF prevention

When PPM is indicated for patients with preserved EF or no pre-existing HF, prophylactic CRT implantation may not be supported, considering the low incidence of PaHF observed in our data (12.9%) and in the two most recent large-scale registry-based studies (10.6% and 25.8%).^21,28^ Instead, cardiac conduction system pacing might be the preferred option over conventional PPM or prophylactic CRT for patients with multiple risk factors of PaHF, particularly for younger patients with a high predicted RV-pacing burden. ^8,40^ According to our data, the absolute mortality rate was higher in older PPM patients; however, the relative contribution of PaHF to mortality gradually increased in younger age groups (P_interaction_<0.001, Figure 2 and Supplementary Figure 6). In elderly PPM patients, comorbidities would exert a heavier impact on mortality than the RV-pacing burden. In contrast, the prognosis of younger PPM patients might be more strongly affected by the RV-pacing burden because they are likely to have fewer comorbidities but more RV-pacing burden during a longer and more active life. ***PaHF monitoring***

After PPM implantation, there are no guidelines regarding when or how often patients with PPM should be monitored for PaHF. Tayal et al. suggested a six-month echocardiographic evaluation would likely identify the majority of PaHF patients based on their findings that the PaHF incidence was significantly higher in the early phase (i.e., within six months post-PPM), whereas the risk significantly decreased during the late phase (i.e., beyond six months). ^21^

Indeed, our data supported these results, demonstrating that the risk of PaHF or its related mortality was highest within six months (Figures 3A and 3B). However, no recommendation was provided after six months. According to our data, the risk was unlikely to attenuate thereafter. Particularly, PaHF-associated mortality began to rise up again approximately 5 years after PPM implant. Furthermore, risk factors for PaHF or PaHF-associated mortality, such as advanced age (≥65 years), DM, CKD/ESRD, or AF, could be accumulated with time, sustaining or increasing the risks, even after six months post-implantation. Therefore, long-term and regular follow-ups of cardiac function may be required after PPM implantation.

### PaHF management

After a PaHF diagnosis, it is also unclear whether CRT-upgrade should be performed immediately or HF medications should be prescribed for a certain period, for example, at least three to six months before CRT-upgrade. In a recent interesting report from Duke University, approximately 60% of patients with left bundle branch block and reduced EF (<35%), exhibited no improvement or even worsening of their LV EF despite three to six months of guideline-directed medical therapy (GDMT). Furthermore, the mean LV EF increase was only 2.0%, significantly smaller than that in patients with a narrow QRS (8.0%, P <0.0001). The authors called into question current guidelines that mandate at least three months of GDMT before CRT implantation, particularly in HF patients with left bundle branch block. ^33^ Likewise, the beneficial effect of pre-CRT GDMT was not clear in pacing-dependent HF patients in some studies. ^7,32^ Indeed, in our study, beneficial effect of MRAs and ACEIs/ARBs were not clear in the secondary cohort of patients with PaHF, while only ARNI and beta-blockers were closely associated with reduced mortality (Table 4). Interestingly, our findings appear to corroborate recent guidelines that recommend the prioritized use of ARNI over ACEIs or ARBs in HF patients with reduced EF. ^41,42^ In addition, the prognosis was significantly better for patients who received CRT-upgrade in combination with medical treatment compared to those who received medical treatment alone without CRT-upgrade. The K-M curve analyses suggest that the prognosis of the two groups began to diverge significantly in the early stages after PaHF diagnosis (Figure 4). Accordingly, immediate changes to biventricular pacing or conduction system pacing along with HF medications, preferably ARNI and beta-blockers, might be a more reasonable option than the delayed upgrade strategy.

The presence of hypertension was also shown to be associated with reduced mortality in the PaHF cohort (Table 5). However, we speculate that this counterintuitive finding is not due to the beneficial effect of hypertension, but rather to the facts that patients with hypertension received HF medications with mortality benefit, especially RAS inhibitors and beta-blockers, much more frequently than those without (absolute SMDs 1.621 and 1.042 for RAS inhibitors and beta-blockers, respectively, Supplementary table 10).

### Limitations

We acknowledge several limitations. First, echocardiographic data were not available. Therefore, we could not separately analyze the incidence of PaHF with preserved or reduced EF. However, our sensitivity analysis used the broad PaHF definition, which likely encompassed PaHF patients with preserved EF and demonstrated consistent results with the main outcomes. Second, device interrogation data were lacking for this cohort. Therefore, the correlation between the RV-pacing burden and the risk of PaHF occurrence or associated mortality could not be evaluated quantitatively. In addition, the lack of RV-pacing data raises the question of whether our PaHF cases were caused by RV-pacing-induced electromechanical dyssynchrony. However, in Korea, CRT-upgrades are refundable only when RV-pacing percentage exceeds 40% on PPM interrogation. Moreover, improved survival of PaHF patients after CRT-upgrade is also likely to support that our PaHF patients were really under the detrimental effects of chronic RV-pacing. Next, it may be premature to conclude ARNI is superior to ACEIs/ARBs for the management of patients with PaHF. More data are needed to validate our findings on the efficacy of ARNI in comparison with ACEIs/ARBs. Finally, the impact of sodium-glucose cotransporter-2 inhibitors on overall mortality was not evaluated due to the limited number of PaHF patients treated with the agent. Further studies are worth conducting regarding the efficacy of sodium-glucose cotransporter-2 inhibitors or other novel HF agents.

However, our study, despite its limitations, may offer valuable insights into the risk profile of PaHF-associated morality over the entire follow-up period, the optimal timing for CRT-upgrade, and the selection of more suitable HF medications for patients with PaHF.

## Conclusions

Our nationwide real-world cohort-based study found that PaHF development was closely associated with increased mortality following PPM implantation while upgrading to CRT devices and treatment with beta-blockers and ARNI were strong protective factors for all-cause death in patients with PaHF. Furthermore, the PaHF incidence and PaHF-associated mortality were highest during the early post-PPM phase (e.g., within six months), but the risks of PaHF occurrence and associated mortality were likely to continue or increase again with time. Therefore, regular and ongoing cardiac function assessments may be required following PPM implantation. In addition, once PaHF is detected, immediate changes into cardiac physiological pacing modalities, such as biventricular pacemaker or cardiac conduction system pacing, must be considered, along with optimal HF medications.

## Data Availability

Data sharing from the Authors is not possible because of legislation from the Korean government. However, additional data are available through approval and oversight by the Korean National Health Insurance Service

## Abbreviations

ACEI: angiotensin-converting-enzyme inhibitors
AF: atrial fibrillation
ARB: angiotensin II receptor blockers
ARNI: angiotensin receptor neprilysin inhibitor
AVB: atrioventricular block
CCI: Charlson comorbidity index
CKD: chronic kidney disease
CRT: cardiac resynchronization therapy
DM: diabetes mellitus
EF: ejection fraction
ESRD: end-stage renal disease
MRA: mineralocorticoid receptor antagonists
PaHF: pacemaker-associated
HF PPM: Permanent pacemaker
SND: sinus node dysfunction

## Acknowledgement

We thank Mi Yang (Seoul Mental Health Welfare Center) for her great help in retrieving the raw data, and Professor Minsu Park (Department of Information and Statistics, Chungnam National University) for his expert opinions on the study design in the early stage of this study.

**Supplementary Figure Legends Supplementary Figure 1. Study profile**

Abbreviations: PPM, permanent pacemaker; HF, heart failure; PaHF, pacemaker-associated heart failure; CRT, cardiac resynchronisation therapy. *, 4 patients died on the day of PaHF diagnosis.

**Supplementary Figure 2. Graphical depiction of immortal-time bias in the PPM cohort**

Abbreviations: PaHF, pacemaker-associated heart failure; PPM, permanent pacemaker.

**Supplementary Figure 3. Cumulative incidence rates of PaHF development according to strict and broad definitions.**

Abbreviations: PaHF, pacemaker-associated heart failure.

**Supplementary Figure 4. Extended Kaplan–Meier survival curves for all-cause death in PPM cohort according to the occurrence of broadly-defined PaHF.**

Abbreviations: PaHF, pacemaker-associated heart failure.

**Supplementary Figure 5. Time-varying hazard ratio of broadly-defined PaHF on all-cause mortality throughout entire follow-up period.**

Hazard ratios across time were estimated using a multivariable Cox regression analysis with a cubic spline function. Solid and dashed lines refer to hazard ratio and its 95% confidence interval, respectively.

Abbreviations: PaHF, pacemaker-associated heart failure; PPM, permanent pacemaker.

**Supplementary Figure 6. Forest plot of association between broadly-defined PaHF and all-cause mortality by subgroups.**

Abbreviations: PaHF, pacemaker-associated heart failure; CI, confidence interval; CAD, coronary artery disease; PAD, peripheral artery disease; CKD, chronic kidney disease; ESRD, end-stage renal disease; VHD, valvular heart disease; AF, atrial fibrillation; COPD, chronic obstructive pulmonary disease; SND, sinus node dysfunction; AVB, atrioventricular block; ACEI, angiotensin-converting enzyme inhibitor; ARB, angiotensin receptor blocker; MRA, mineralocorticoid receptor antagonist.

**Supplementary Figure 7. Graphical depiction of immortal-time bias in the PaHF cohort** Abbreviations: CRT, cardiac resynchronization therapy; PaHF, pacemaker-associated heart failure.

**Supplementary Figure 8. Extended Kaplan–Meier survival curves for all-cause death in PaHF cohort.**

Overall survival according to CRT-upgrade after adjusting for immortal-time bias and potential confounders (A) before propensity score matching, and (B) after propensity score matching. Abbreviations: CRT, cardiac resynchronization therapy; PaHF, pacemaker-associated heart failure.

